# Socioeconomic modification of the diet quality-HbA1c association among U.S. adults: A survey-weighted interaction analysis

**DOI:** 10.64898/2026.05.04.26352401

**Authors:** Eugene Ampofo, Charles Apprey, Mary Amoako, Fiifi Derrick Turkson

**Author notes:** **Corresponding author Email:** (EA). **Acknowledgments** The authors thank the staff and participants of the National Health and Nutrition Examination Survey for providing the public datasets that made this research possible. **Competing Interests** The authors have declared that no competing interests exist.

## Abstract

Traditional nutrition science often proceeds under the assumption of a universal metabolic return to healthy eating, yet social environments may fundamentally modify these biological associations. This investigation utilized survey-weighted data from the National Health and Nutrition Examination Survey (NHANES 2017–2023) representing a weighted population of 286 million adults aged 20 years and older to test for association heterogeneity in the diet-glycemia relationship. Dietary exposure was operationalized as energy-adjusted nutrient density scores derived via the residual method to measure the healthfulness of intake independent of total caloric volume. The primary outcome was HPLC-measured glycated hemoglobin (HbA1c) modeled as a continuous variable. Multivariable interaction models evaluated the Income-to-Poverty Ratio (PIR) as a formal effect modifier, adjusting for age, sex, race/ethnicity, body mass index, and smoking status. Analysis demonstrated that while quality-weighted nutrient intake levels remained statistically uniform across income tiers (*p*=0.207), multivariable interaction models identified significant modification of the diet-HbA1c association by socioeconomic position (*p*=0.028 for interaction). In the low-income reference group, higher quality nutrient intake was associated with a significant protective decline in HbA1c (*β*=−6.11×10−5 percentage points per kilocalorie, *p*=0.017). Conversely, this protective association was attenuated to non-significance for the middle-income stratum (*β*=3.31×10−6, *p*=0.849). Sensitivity analysis restricted to participants without clinical diabetes showed the interaction was non-significant (*p*=0.859), identifying an observed boundary condition where differential associations are most evident in pathological states. These findings suggest association heterogeneity where identical dietary behaviors relate to divergent glycemic patterning across socioeconomic groups. Socioeconomic position appears to function as an effect modifier of the diet-HbA1c relationship rather than a mere confounder. Such patterns suggest that the metabolic correlates of healthy eating are socially patterned, potentially due to structural factors that constrain the associations expected from improved nutrition.

## 2. INTRODUCTION

Glycated hemoglobin (HbA1c) remains the primary clinical benchmark for tracking three-month glucose averages and predicting long-term diabetic complications [1,2]. More than 38 million adults in the United States currently navigate type 2 diabetes, a condition driven largely by suboptimal nutrient intake [3,4]. Clinical guidelines emphasize high-quality diet as a modifiable defense against metabolic instability [5]. Yet, decades of federal investment in behavioral counseling have failed to close health gaps for marginalized groups [6].

Socioeconomic position determines these outcomes [7,8]. Disadvantaged populations consistently face higher mean HbA1c and greater barriers to effective disease management [1,2]. These disparities stem from structural conditions, including neighborhood food deprivation and the cumulative physiological toll of chronic psychosocial stress [4,9]. Traditional research treats dietary choices and financial status as separate variables, ignoring their potential interaction [10].

A major knowledge gap persists regarding the “universal metabolic return” assumption. Most public health models presume identical biological benefits from healthy eating regardless of social context [10]. If structural disadvantage acts as a physiological attenuator, it may decouple healthy behaviors from their expected metabolic outcomes [11]. Advising marginalized groups to “eat better” is an incomplete equity strategy if the social environment blunts the body’s ability to translate nutrients into improved glycemic control [10,12]. Contemporary literature lacks a population-level test of whether the metabolic association of nutrient quality is formally modified by an individual’s financial standing.

The National Health and Nutrition Examination Survey (NHANES) pairs validated 24-hour dietary recalls with objective HPLC-measured biomarkers [2,6]. This study utilized contemporary data representing approximately 286 million adults (2017–2023) to examine whether socioeconomic position formally modifies the association between diet quality and HbA1c. We tested three specific hypotheses: first, that higher diet quality associates with lower HbA1c; second, that lower-income groups exhibit higher HbA1c levels; and third, that the protective association between diet and glycemia is significantly attenuated among lower socioeconomic groups. Moving beyond “average effects” to the distribution of benefits builds a scientific case for structural interventions in public health nutrition.

## 3. METHODS

### 3.1 Data Integration and Population Snapshot

We merged the 2017–March 2020 prepandemic NHANES file with the 2021–2023 continuous cycles to create a current metabolic snapshot of the United States. NHANES data were accessed for research purposes in January 2026. The authors did not have access to identifiable participant information. NHANES uses a complex, multistage probability design to represent the non-institutionalized civilian population. Participants completed standardized physical examinations and biospecimen collection at Mobile Examination Centers (MECs) following in-home interviews. Reporting followed STROBE guidelines to ensure technical transparency.

### 3.2. Sample Power and Boundaries

The analytic cohort included adults aged 20 years and older. We excluded pregnant or breastfeeding individuals due to their unique metabolic baselines. The final weighted population represents approximately 286 million adults. A weighted sample of this magnitude provides the robust statistical power needed to detect formal interaction effects and association heterogeneity that smaller clinical trials lack the granularity to identify.

### 3.3. Primary Outcome: Continuous HbA1c

HPLC-measured glycated hemoglobin (HbA1c) served as the primary outcome. Whole blood was analyzed at a central laboratory using high-performance liquid chromatography (HPLC), a method known for linear accuracy across various red cell counts. Modeling HbA1c as a continuous variable captures subtle physiological changes that binary diabetes classifications often mask. Objective laboratory values reduce recall bias compared to self-reported status.

### 3.4. Dietary Exposure and Energy Adjustment

Dietary exposure was the mean total nutrient intake of Day 1 and Day 2 24-hour recalls. This averaging reduces within-person variation and provides a more stable estimate of usual intake. Recalls utilized the Automated Multiple-Pass Method (AMPM) to minimize underreporting. We modeled nutrient intake as a data-derived continuous variable rather than comparing it against external USDA benchmarks like the Healthy Eating Index. The residual method decoupled nutrient quality from total energy, creating a calorie-independent quality score.

### 3.5. Socioeconomic Position and Interaction Model

Income-to-Poverty Ratio (PIR) operationalized socioeconomic position. PIR was split into tertiles: Low Income (< 1.3), Middle Income (1.3 to < 3.5), and High Income (≥ 3.5). Testing for association heterogeneity was pre-specified through a formal interaction term (Intake × PIR Tertile). This design directly evaluates the “Unequal Returns” hypothesis: that structural disadvantage blunts the metabolic benefits of a high-quality diet.

### 3.6. Statistical Architecture and Adjustment Strategy

Analyses were executed in R version 4.3.1 using the *survey* and *srvyr* packages. Pooled weights followed NCHS technical guidance for combined cycles. Multivariable models adjusted for sociodemographic and clinical confounders, including chronological age, biological sex, and self-reported race/ethnicity. Models further controlled for Body Mass Index (BMI) and smoking status. Stratified models calculated slope coefficients (*β*) within each income stratum to visualize divergent trajectories.

### 3.7. Missing Data and Sensitivity Analysis

Missing PIR data (~9.9%) were handled via Multiple Imputation by Chained Equations (MICE) with predictive mean matching. This semi-parametric approach preserves the multivariate structure of the dataset. We generated five imputed datasets (*m*=5), executed models across these sets, and combined results via Rubin’s rules. Sensitivity testing repeated interaction models after restricting the sample to non-diabetic participants to determine if social modifiers are most visible once regulatory resilience is compromised.

### 3.8. Ethics Statement

This secondary analysis utilized publicly available, de-identified data and was exempt from Institutional Review Board oversight. NHANES protocols are approved by the NCHS Research Ethics Review Board and participants provided written informed consent. Significance was set at *p*<0.05 using two-sided tests.

## 4. RESULTS

### 4.1 Descriptive Characteristics and Socioeconomic Distribution

United States adults (weighted N = 286,698,034) maintain energy-adjusted nutrient intake quality levels that are statistically uniform across income-to-poverty ratio (PIR) tiers (*p*=0.207). In contrast, mean glycated hemoglobin (HbA1c) levels follow a linear financial gradient (*p*=0.003), with the highest levels recorded in the low-income group (mean: 5.70%, SD: 1.11) and the lowest in the high-income group (mean: 5.56%, SD: 0.81). As detailed in Table 1, middle-income adults recorded the highest mean Body Mass Index (BMI) at 28.14 kg/m^2^ (*p*=0.005). Demographic distribution shifted across PIR tiers; high-income participants were older (mean: 41.4 years) and predominantly Non-Hispanic White (75.2%), while the low-income group was younger (mean: 31.5 years) and carried a higher proportion of Mexican American (17.4%) and Non-Hispanic Black (20.9%) participants (*p*<0.001).

**Table 1.**
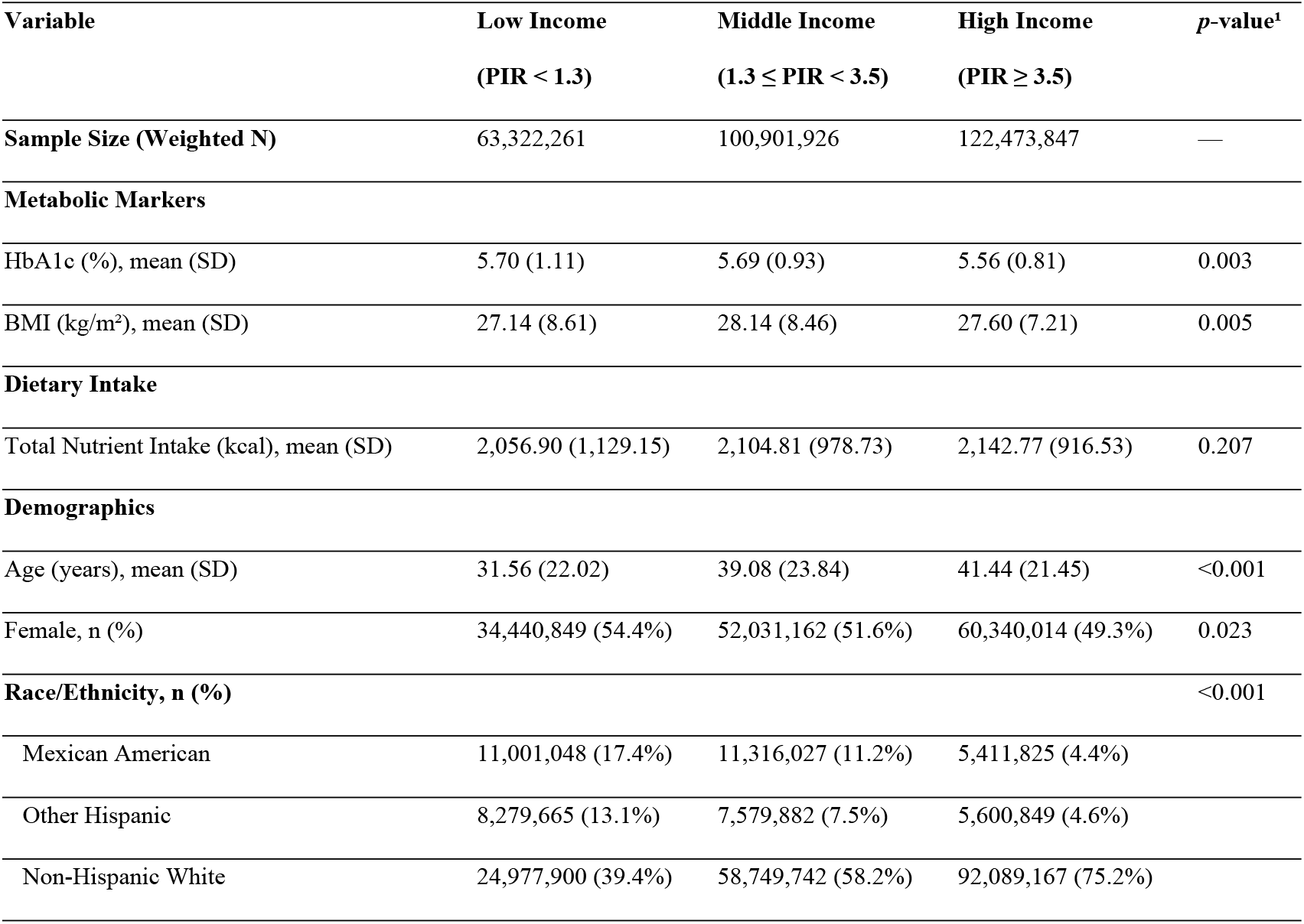
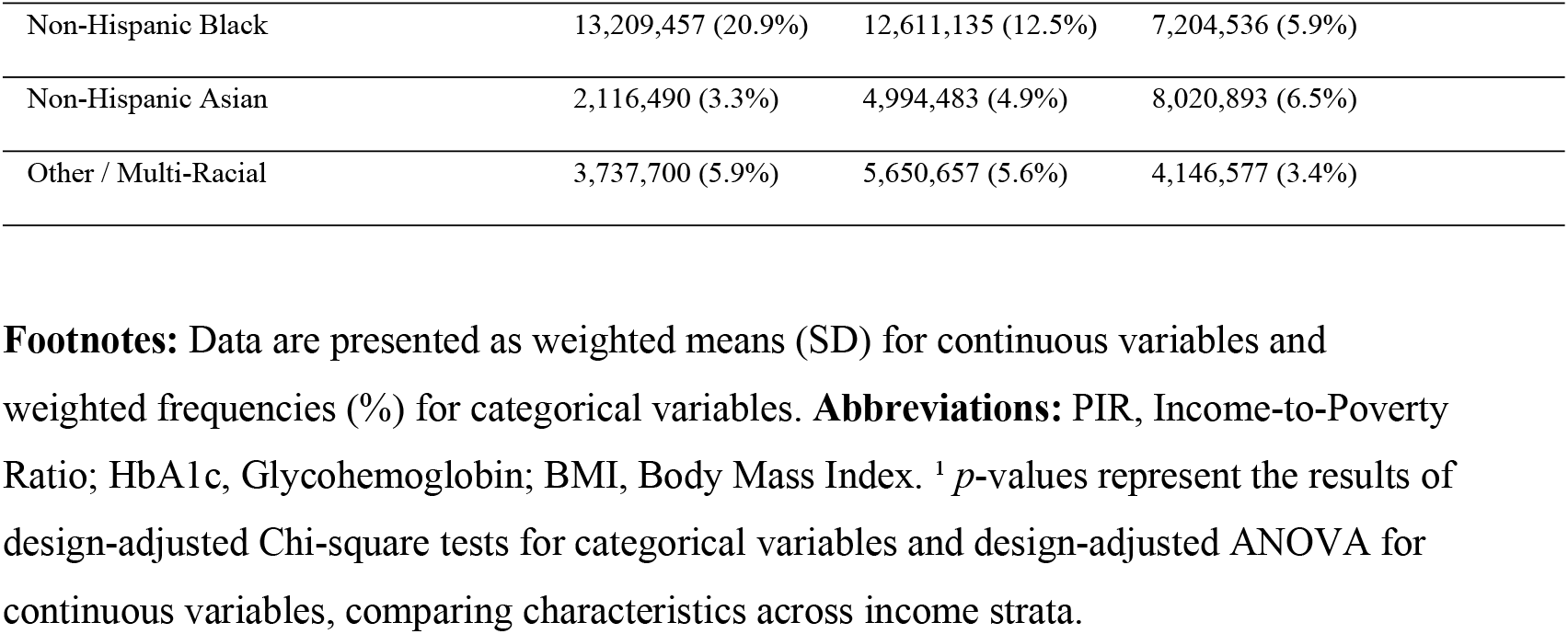
Weighted Baseline Characteristics of U.S. Adults Stratified by Income-to-Poverty Ratio (PIR)

**Table 2.**
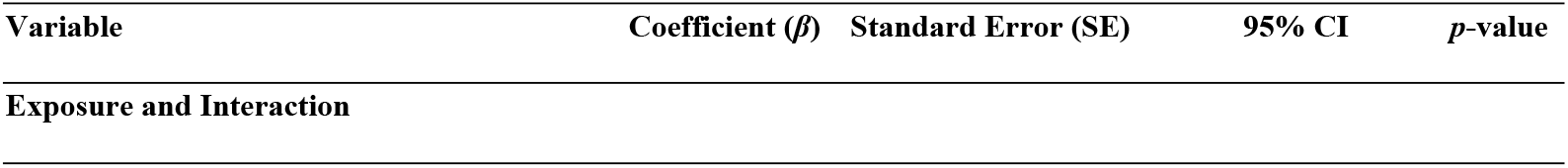
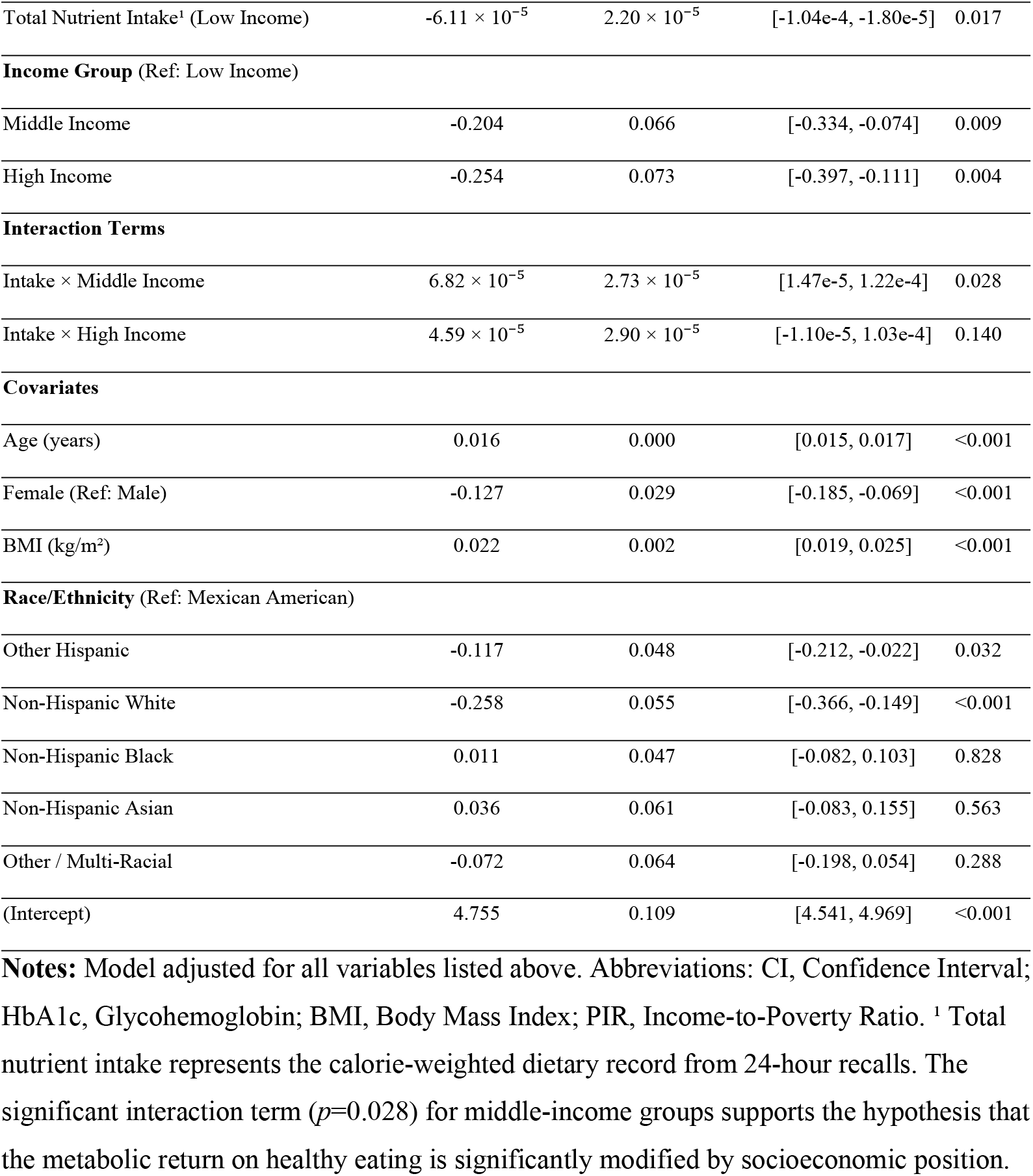
Weighted multivariable regression analysis of the interaction between nutrient intake and socioeconomic position on HbA1c (%).

| Variable | Coefficient ($\beta$) | Standard Error (SE) | 95% CI | *p*-value |
| --- | --- | --- | --- | --- |
| Exposure and Interaction |  |  |  |  |
| Total Nutrient Intake<sup>1</sup> (Low Income) | $-6.11 \times 10^{-5}$ | $2.20 \times 10^{-5}$ | [-1.04e-4, -1.80e-5] | 0.017 |
| **Income Group** (Ref: Low Income) |  |  |  |  |
| Middle Income | -0.204 | 0.066 | [-0.334, -0.074] | 0.009 |
| High Income | -0.254 | 0.073 | [-0.397, -0.111] | 0.004 |
| **Interaction Terms** |  |  |  |  |
| Intake × Middle Income | $6.82 \times 10^{-5}$ | $2.73 \times 10^{-5}$ | [1.47e-5, 1.22e-4] | 0.028 |
| Intake × High Income | $4.59 \times 10^{-5}$ | $2.90 \times 10^{-5}$ | [-1.10e-5, 1.03e-4] | 0.140 |
| **Covariates** |  |  |  |  |
| Age (years) | 0.016 | 0.000 | [0.015, 0.017] | <0.001 |
| Female (Ref: Male) | -0.127 | 0.029 | [-0.185, -0.069] | <0.001 |
| BMI (kg/m<sup>2</sup>) | 0.022 | 0.002 | [0.019, 0.025] | <0.001 |
| **Race/Ethnicity** (Ref: Mexican American) |  |  |  |  |
| Other Hispanic | -0.117 | 0.048 | [-0.212, -0.022] | 0.032 |
| Non-Hispanic White | -0.258 | 0.055 | [-0.366, -0.149] | <0.001 |
| Non-Hispanic Black | 0.011 | 0.047 | [-0.082, 0.103] | 0.828 |
| Non-Hispanic Asian | 0.036 | 0.061 | [-0.083, 0.155] | 0.563 |
| Other / Multi-Racial | -0.072 | 0.064 | [-0.198, 0.054] | 0.288 |
| (Intercept) | 4.755 | 0.109 | [4.541, 4.969] | <0.001 |

**Notes:** Model adjusted for all variables listed above. Abbreviations: CI, Confidence Interval; HbA1c, Glycohemoglobin; BMI, Body Mass Index; PIR, Income-to-Poverty Ratio. <sup>1</sup> Total nutrient intake represents the calorie-weighted dietary record from 24-hour recalls. The significant interaction term ( $p=0.028$ ) for middle-income groups supports the hypothesis that the metabolic return on healthy eating is significantly modified by socioeconomic position.

### 4.2. Multivariable Interaction Modeling

Multivariable linear regression identifies socioeconomic position (SEP) as a formal modifier of the association between quality-weighted nutrient intake and HbA1c (*p*=0.028 for interaction). For the low-income reference group (PIR < 1.3), higher nutrient quality associates with a protective decline in HbA1c (*β*=−6.11×10−5, *p*=0.017). This association is significantly modified in the middle-income tier (1.3≤*PIR*<3.5), where the interaction term is positive (*β*=6.82×10−5, *p*=0.028). The interaction term for the high-income group did not reach statistical significance (*β*=4.59×10-5, *p*=0.140). Age (*β*=0.016), BMI (*β*=0.022), and female sex (*β*=-0.127) remained robust independent predictors of glycemia across all models (*p*<0.001). As conceptualized in Figure 1: Conceptual Framework of Unequal Returns, these results imply that structural barriers such as chronic psychosocial stress or neighborhood food environments likely act as physiological attenuators. While age and BMI remain aggressive independent drivers of glycemia (*p*<0.001), they do not erase this significant social interaction.

**Fig 1.**
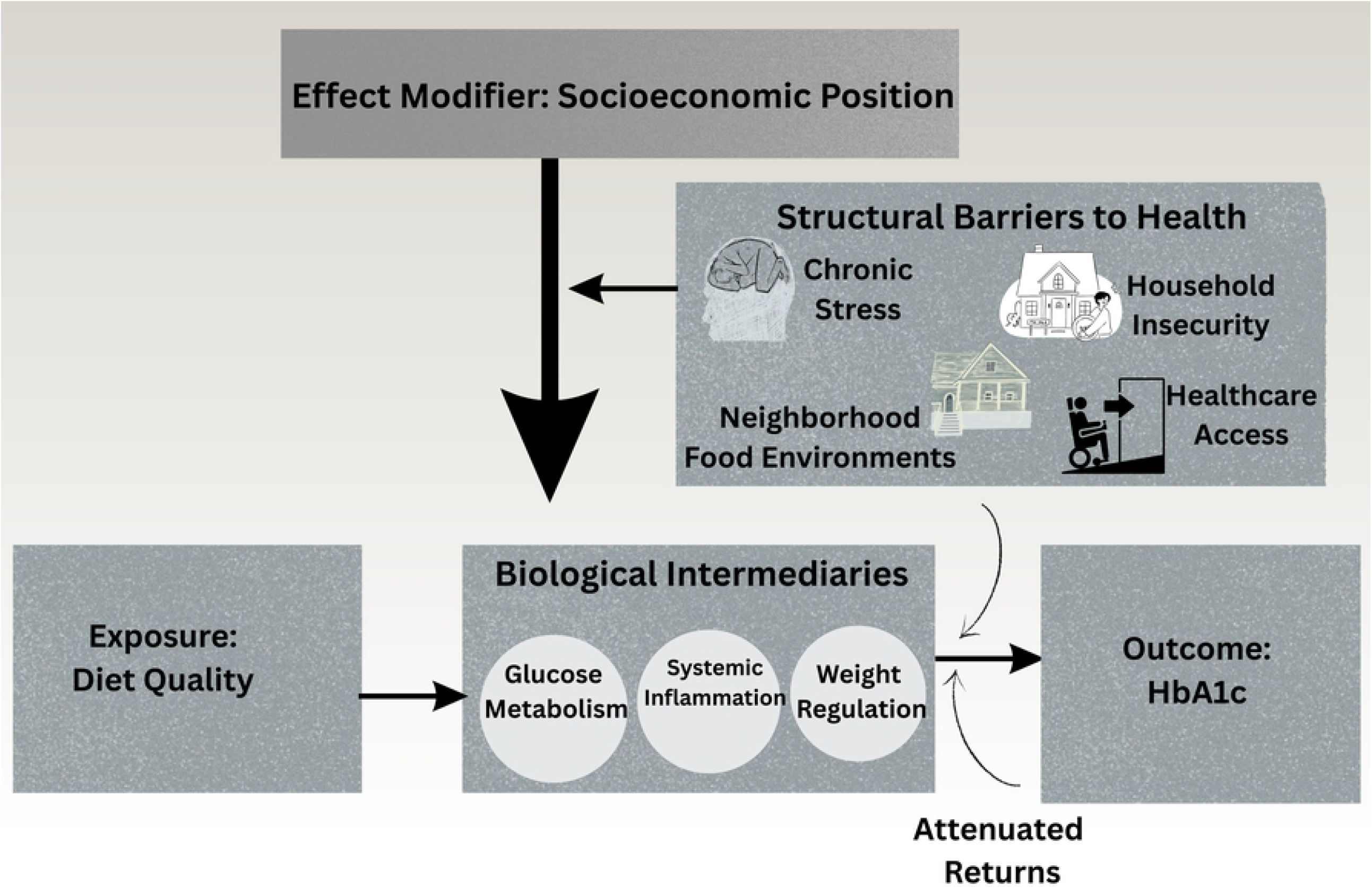
Conceptual framework of unequal metabolic returns to healthy eating. This diagram illustrates the hypothesized pathway where diet quality (exposure) influences glycated hemoglobin (outcome) through biological intermediaries such as glucose metabolism, systemic inflammation, and weight regulation. Socioeconomic position acts as a formal effect modifier, where structural barriers including chronic stress, household insecurity, and neighborhood food environments attenuate the physiological returns of healthy eating in certain populations.

### 4.3 Stratified Association Estimates

Stratified slope coefficients indicate divergent metabolic associations across income groups. The protective association between diet and glycemia was highest for the low-income stratum (*β*=−5.78×10−5, *p*=0.053), though it reached only marginal statistical significance. This association flattens to a non-significant horizontal line for middle-income adults (*β*=3.31×10−6, *p*=0.849). High-income adults exhibit a modest, non-significant protective association (*β*=−1.31×10−5, *p*=0.619) (Liang et al., 2020; Table 3). Figure 2 provides a visual representation of these divergent trajectories, showing a decline in predicted HbA1c for the low-income group compared to the flat trajectory observed in the middle-income tier.

**Table 3.** Stratified Associations between Nutrient Intake and HbA1c (%) by Income Group.

| Income Stratum | Coefficient ($\beta$)* | Standard Error (SE) | 95% Confidence Interval (CI) | p-value |
| --- | --- | --- | --- | --- |
| Low Income (PIR < 1.3) | $-5.78 \times 10^{-5}$ | $2.77 \times 10^{-5}$ | $[-1.16 \times 10^{-4}, 8.67 \times 10^{-7}]$ | 0.053 |
| Middle Income ($1.3 \leq \text{PIR} < 3.5$) | $3.31 \times 10^{-6}$ | $1.71 \times 10^{-5}$ | $[-3.29 \times 10^{-5}, 3.95 \times 10^{-5}]$ | 0.849 |
| High Income (PIR $\geq 3.5$) | $-1.31 \times 10^{-5}$ | $2.57 \times 10^{-5}$ | $[-6.76 \times 10^{-5}, 4.15 \times 10^{-5}]$ | 0.619 |

**Footnotes:** \*Coefficient ( $\beta$ ) represents the change in HbA1c (%) per unit increase in total nutrient intake (kcal), adjusted for age, sex, race/ethnicity, and BMI. **Abbreviations:** PIR, Income-to-Poverty Ratio; CI, Confidence Interval; HbA1c, Glycohemoglobin.

**Fig 2.**
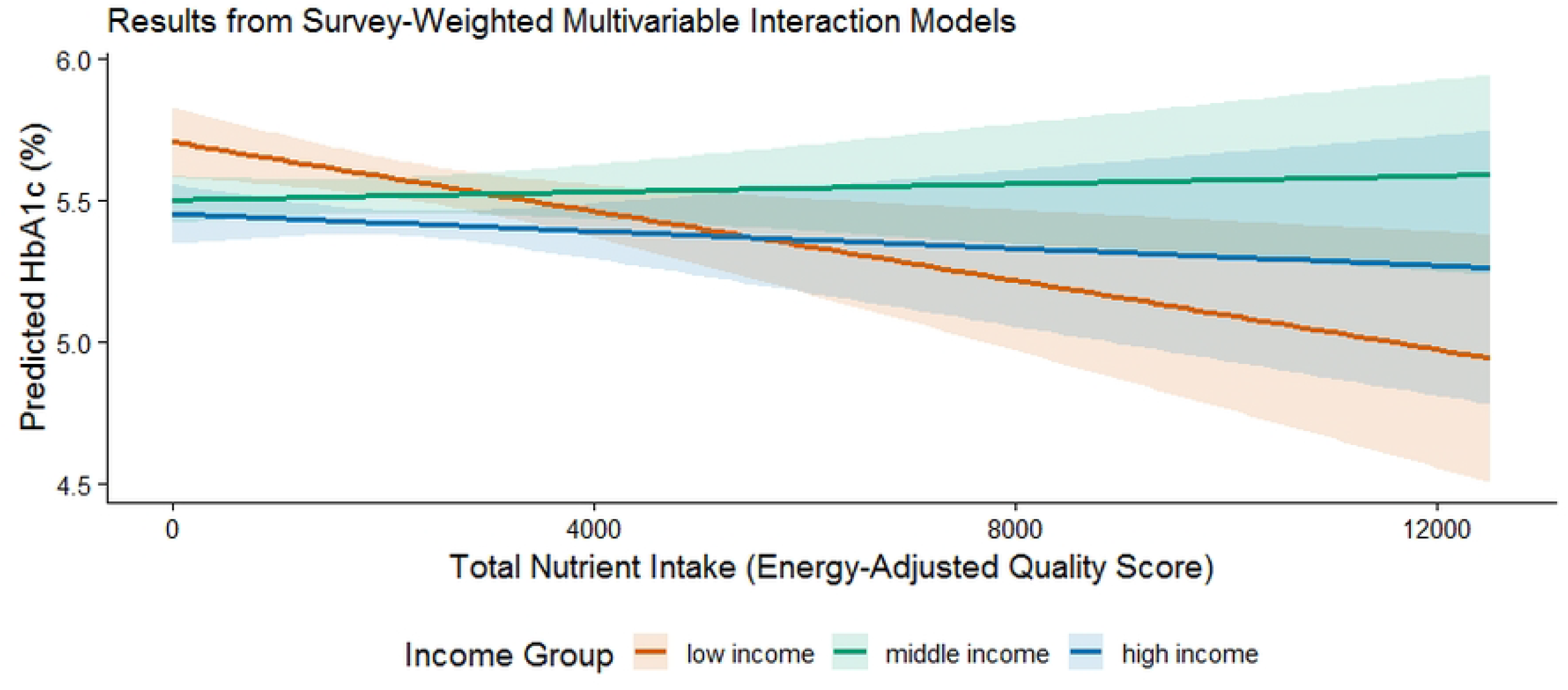
Predicted metabolic returns to healthy eating by income group. Results from survey-weighted multivariable interaction models depicting predicted glycated hemoglobin (%) levels relative to calorie-weighted nutrient intake quality. Slopes are stratified by income-to-poverty ratio (PIR) tiers: low income (PIR < 1.3), middle income (1.3 ≤ PIR < 3.5), and high income (PIR ≥ 3.5). The visualization demonstrates the significant attenuation of the protective diet-HbA1c association specifically within the middle-income stratum.

### 4.4 Sensitivity Analysis and Boundary Conditions

Sensitivity testing in Table 4 restricted the sample to non-diabetic participants to identify potential boundary conditions for the observed interaction. In this subsample, the interaction terms for both middle-income (*p*=0.859) and high-income (*p*=0.917) groups were non-significant. The direct association between nutrient intake and HbA1c was also attenuated and non-significant in the non-diabetic population (*β*=1.68×10−5, *p*=0.300). Age (*β*=0.009), BMI (*β*=0.013), and Non-Hispanic White race (*β*=−0.135) remained consistent and significant predictors of HbA1c levels within this subsample (*p*<0.001). The model intercept for non-diabetic participants was recorded at 4.755 (*p*<0.001).

**Table 4.** Sensitivity Analysis: Weighted Multivariable Regression of the Interaction between Nutrient Intake and Income among Non-Diabetic Participants.

| Variable | Coefficient ($\beta$) | 95% CI | p-value |
| --- | --- | --- | --- |
| **Exposure and Interaction** |  |  |  |
| Total Nutrient Intake<sup>1</sup> (Low Income) | $1.68 \times 10^{-5}$ | [-1.70e-5, 5.06e-5] | 0.300 |
| **Income Group (Ref: Low Income)** |  |  |  |
| Middle Income | -0.007 | [-0.109, 0.096] | 0.892 |
| High Income | -0.064 | [-0.158, 0.031] | 0.168 |
| **Interaction Terms** |  |  |  |
| Intake × Middle Income | $-3.79 \times 10^{-6}$ | [-4.92e-5, 4.16e-5] | 0.859 |
| Intake × High Income | $-2.01 \times 10^{-6}$ | [-4.32e-5, 3.92e-5] | 0.917 |
| **Key Covariates** |  |  |  |
| Age (years) | 0.009 | [0.008, 0.010] | < 0.001 |
| BMI (kg/m<sup>2</sup>) | 0.013 | [0.010, 0.016] | < 0.001 |
| Non-Hispanic White (Ref: Mex. Amer.) | -0.135 | [-0.201, -0.069] | < 0.001 |

**Notes:** Model adjusted for sex and all race/ethnicity categories (not all shown for brevity). <sup>1</sup> Total nutrient intake represents calorie-weighted dietary record. **Abbreviations:** CI, Confidence Interval; HbA1c, Glycohemoglobin; PIR, Income-to-Poverty Ratio.

## 5. DISCUSSION

### 5.1 Findings and the Metabolic Returns Paradox

U.S. adults maintain statistically uniform nutrient intake profiles across income tiers, yet their glycemic outcomes follow a rigid socioeconomic ladder. This discrepancy identifies a breakdown in the assumed biological payoff of nutrition. For middle-income adults, the protective association between high-quality nutrient intake and glycated hemoglobin is effectively absent, contrasting with the significant protective return observed in lower-income peers. These divergent trajectories suggest that structural factors correlate with a blunted metabolic association with healthy eating in specific segments of the population [7, 8].

### 5.2 Interpretation and the MDR Framework

The “blunting” of dietary benefits among middle-income groups aligns with the Minorities’ Diminished Return (MDR) framework, which suggests that social resources do not yield equal health protection for all populations [10, 11]. While previous work establish that educational attainment relates to greater fruit and vegetable intake for White but not Black adults [10], our investigation identifies a similar phenomenon regarding the physiological translation of nutrients into glycemic control. The data challenge the assumption of biological uniformity, identifying a “metabolic stasis” where increasing nutrient density fails to shift the HbA1c needle for specific socioeconomic groups [7].

### 5.3 Potential Mechanisms: Physiological Attenuators

Structural conditions likely act as physiological attenuators that decouple healthy behaviors from their expected metabolic outcomes [10,13]. High allostatic load, the cumulative cost of adapting to chronic stressors like housing instability or material deprivation is linked to impaired glucose metabolism independently of caloric volume [8,14]. Our sensitivity analysis suggests these social modifiers become most visible once metabolic regulation is already compromised by clinical pathology. In metabolically healthy individuals, the body appears to maintain enough regulatory resilience to process nutrients effectively across all social strata [15].

### 5.4 Implications for Clinical Practice

Clinical models often presume a universal response to nutritional counseling. Our data suggest that a patient’s lack of glycemic response to a high-quality diet may stem from environmental constraints rather than individual non-compliance. Assessment of social determinants of health, such as medication affordability and food security, provides necessary context for clinical outcomes [4, 16]. Clinical guidance should account for the possibility that the metabolic benefits of nutrition are constrained by deep-seated structural factors that individual behavioral change cannot always overcome.

### 5.5. Directions for Future Research

Longitudinal studies are essential to establish a temporal sequence and determine if structural disadvantage causally blunts the metabolic response to nutrition over time. Future work should utilize mediation analyses to identify if specific factors, such as sleep disruption or out-of-pocket medication costs, drive these divergent returns. There is a critical need to test whether “Food as Medicine” interventions yield different metabolic returns across socioeconomic groups to ensure interventions do not inadvertently widen health gaps.

### 5.6 Strengths and Methodological Shortcomings

Relying on HPLC-measured HbA1c eliminates the recall bias inherent in self-reported diabetes status. The large weighted sample size of 286 million adults provides the statistical power necessary to detect interaction effects that smaller trials lack the granularity to identify. However, the cross-sectional nature of NHANES prevents establishing a causal link between dietary shifts and metabolic change. 24-hour dietary recalls are susceptible to random measurement error, though using the mean of multiple days improves precision. Finally, the Income-to-Poverty Ratio may not capture the full complexity of household wealth or neighborhood food nuances.

### 5.7. Conclusions

Healthy eating is associated with unequal metabolic returns across U.S. income groups. While dietary guidelines are uniform, the biological association between nutrient intake and glycemia is socially patterned. This biological blunting suggests that structural inequity may act as a physiological blocker to metabolic improvement. Narrowing the glycemic equity gap involves moving beyond behavior-only models to address the environmental blockers of metabolic health.

## Data Availability

No data was generated by this study. The following existing data sources were used: NHANES database available via https://www.cdc.gov/nchs/nhanes/.

**Acknowledgments**

The authors thank the participants and staff of the National Health and Nutrition Examination Survey (NHANES) for their role in providing the publicly available datasets that made this research possible.

